# Systems and organisational change to advance gender equity in healthcare leadership: a mixed-methods protocol

**DOI:** 10.1101/2025.10.12.25337851

**Authors:** Belinda Garth, Anusha Ramani-Chander, Jenny Proimos, Darren Rajit, Erwin Loh, Elizabeth Sigston, Graeme Currie, Kathleen Riach, Mariam Mousa, Helena Teede, the Advancing Women in Healthcare Leadership investigator group

## Abstract

**Background:** Despite comprising over 70% of the global healthcare workforce, women remain significantly underrepresented in healthcare leadership. Structural and systemic barriers persist across academic medicine, health services, and professional organisations, limiting career progression and leadership opportunities for women. Existing efforts often focus on individual-level interventions, overlooking the broader organisational and systemic contexts that shape leadership pathways. Urgent, coordinated action is needed to address gender inequity through sustainable, evidence-informed systems change.

**Methods:** This protocol outlines the Organisation Change Management (OCM) workstream within the Australian Advancing Women in Healthcare Leadership (AWHL) initiative—a nationally implemented, multi-sector partnership. The initiative applies a mixed-methods, coproduction approach to implement and evaluate multi-level interventions aimed at advancing gender equity in healthcare leadership. Guided by the Consolidated Framework for Implementation Research, the Learning Health System framework, and the Reach, Effectiveness, Adoption, Implementation and Maintenance evaluation model, the study engages stakeholders across outer (policy, regulation, funding) and inner (organisational culture, leadership structures) settings to drive systemic and organisational changes to enhance gender equity in leadership. Data collection includes administrative datasets and policy documents, semi-structured interviews, and surveys across partner organisations. Findings will inform tailored interventions and an implementation toolkit, developed and evaluated through iterative stakeholder engagement.

**Discussion:** This is the first national initiative to apply a systems-level, coproduced approach to gender equity in healthcare leadership, engaging strategic partners including health services, professional colleges and associations, government, and women in the workforce. By leveraging implementation science and systems change methodologies, the initiative aims to accelerate sustainable organisational transformation. The protocol provides a replicable framework for advancing equity in healthcare leadership and beyond.

## BACKGROUND

Women comprise 70% of the global healthcare workforce, yet remain significantly underrepresented in healthcare leadership (1,2). Despite international policy mandates prioritising gender equality – such as the United Nations Sustainable Development Goal Five ‘Achieve Gender Equality and Empower All Women and Girls’, World Health Organization objectives, and the Lancet Challenge (3–5) – progress has been slow. Globally, women hold only 31% of executive director positions and 20% of board chair roles in global health organisations (1), and just 28% of Dean positions in leading public health and medical schools (1,6). In academic medicine, women are less likely than men to be promoted to associate or full professor, or appointed to department chair roles (7). Persistent income disparities further compound these inequities (8). Promotion and tenure processes have been identified as key contributors to gender disparities in career advancement (9). These inequities are well documented, including across public and private health services (10), global health institutions (11), academic medicine (12), and leadership within medical colleges and specialty societies (13). The intersection of gender with race and ethnicity further compounds inequity and marginalises many women from progressing to leadership (14,15).

Globally, progress toward gender equity in workplace leadership remains slow across all sectors. At the current rate of change, achieving equal representation is projected to take another 140 years (16). While this estimate is not specific to healthcare, it highlights the urgent need for coordinated action, particularly in a sector such as healthcare where women comprise the majority of the workforce. Women in healthcare should not have to wait generations for equitable leadership opportunities. This persistent inequality is not only unjust; it demands immediate and sustained action. The question is no longer whether gender equity in healthcare leadership is necessary, or why it is essential, but *how it can be achieved* through sustainable, system-wide transformation.

Diverse leadership benefits everyone. In workplaces, diverse leadership is a key driver for economic growth and workforce empowerment. It fosters a more motivated and engaged workforce, reduces attrition, enhances job satisfaction, and contributes to strategic advantages across social and economic development (1). Organisations with diverse leadership consistently demonstrate improved performance, productivity, and profitability (17). In healthcare, increasing the representation of women in leadership is associated with improved quality of care, lower patient mortality rates, and more equitable health outcomes for patients, particularly for women and children (1,11,18).

Despite the well-established benefits of diverse leadership, structural and systemic barriers to women’s advancement in healthcare leadership persist (19–22). Progress has been limited, in part, by a historical focus on organisational norms that prioritise individual-level solutions – often placing the onus on women themselves – rather than addressing the broader organisational and systemic contexts in which they work. To move forward, there is a recognised need to shift the responsibility for change from individuals to collective, system-wide and organisational action (23,24). Meaningful and lasting progress requires coordinated, sector-wide strategies that are evidence-based and replace fragmented, adhoc efforts. This involves active engagement, partnership, and coproduction with key stakeholders, drawing on cross-sector expertise and centring the voices of women in the workforce (20,25).

### The Advancing Women in Healthcare Leadership (AWHL) initiative

It is in this context that the Australian, nationally implemented AWHL initiative was established. AWHL aims to partner, coproduce, implement, evaluate and scale multi-faceted system, organisational and individual level interventions that measurably improve career progression to leadership for women in healthcare (26). A stakeholder matrix guided the establishment of this national partnership, which spans leading health services, professional colleges and associations, and government. The process was guided by cross-sector and international academic expertise. Crucially, the voices of women have been recognised as a collective stakeholder, ensuring that interventions are tailored and contextualised to reflect their needs, preferences, and lived experiences; ultimately informing more effective implementation.

### Organisation Change Management (OCM) Workstream

The Organisational Change Management (OCM) workstream within the AWHL initiative focuses on transforming the systems and organisations in which individuals train and work.

Its objective is to embed equity into the culture of healthcare systems, ensuring that change is systemic, collective, and sustainable.

The OCM workstream:

- *Aligns individual advancement with systemic reform:* It aims to ensure that women’s progression into leadership is not reliant on exceptional personal effort or resilience, but is instead supported by equitable policies, practices, and institutional norms.
- *Positions equity as an organisational responsibility:* Rather than placing the burden on women to adapt to existing structures, OCM reorients those structures to become more inclusive, adaptive, and reflective of the diverse healthcare workforce.

The overarching purpose of this workstream is to coproduce evidence-based, sustainable organisational and systems-level change – partnering with women in the workforce, researchers, clinicians and leaders – to advance women in healthcare leadership. The specific objectives are to:

1. Identify evidence-based drivers of sustainable complex systems change for gender equity in healthcare leadership, with particular focus on the systems and organisations in which individuals train and work.
2. Coproduce, implement and evaluate evidence-based systemic and organisational changes to enhance gender equity in leadership.

## METHODS AND ANALYSIS

### Background and coproduction approach

A multiple-case study design will be used, with the partner organisation as the unit of analysis (27). Partner organisations have been selected because they represent strategic stakeholders across health services, professional colleges and associations, government, and women in the healthcare workforce. This design enables in-depth exploration of each case within its real-world context, while facilitating cross-case learning.

The AWHL initiative employs coproduction-implementation research methods, engaging partner organisations throughout each stage (26,28,29). Strong stakeholder partnerships are foundational to this approach (30). While the definition of coproduction varies, in AWHL it is understood as: stakeholders working together from the outset with shared power and equal partnership, bringing different perspectives including their local context to influence the way interventions are designed, implemented and delivered, to optimise impact on policy and practice (31,32). Importantly, solutions are created with active input from those traditionally considered ‘decision-makers’ (e.g. government, member organisations and health service managers) and those traditionally considered on the receiving-end or ‘subjected’ to these solutions (e.g. women in the workforce) (30). The data collection and analysis process is designed to ensure equitable representation of voices and contributions from individuals across varying positions of power.

This approach facilitates collective action through the construction of a shared agenda (28), activities and actions to implement meaningful, evidence-based and actionable change-focused organisational interventions for measurable impact. This then leads into collaborative implementation of tailored solutions, led by partners and facilitated by academic experts.

### Implementation frameworks

Our approach will utilise what we term the Monash Systems Change Approach, which incorporates three existing frameworks to drive evidence-based, complex systemic and organisational change for gender equity in healthcare leadership: 1) the Consolidated Framework for Implementation Research (CFIR), 2) the Learning Health System (LHS), and 3) the Reach, Effectiveness, Adoption, Implementation and Maintenance (RE-AIM) evaluation framework, as shown in Figure 1.

**Figure 1.**
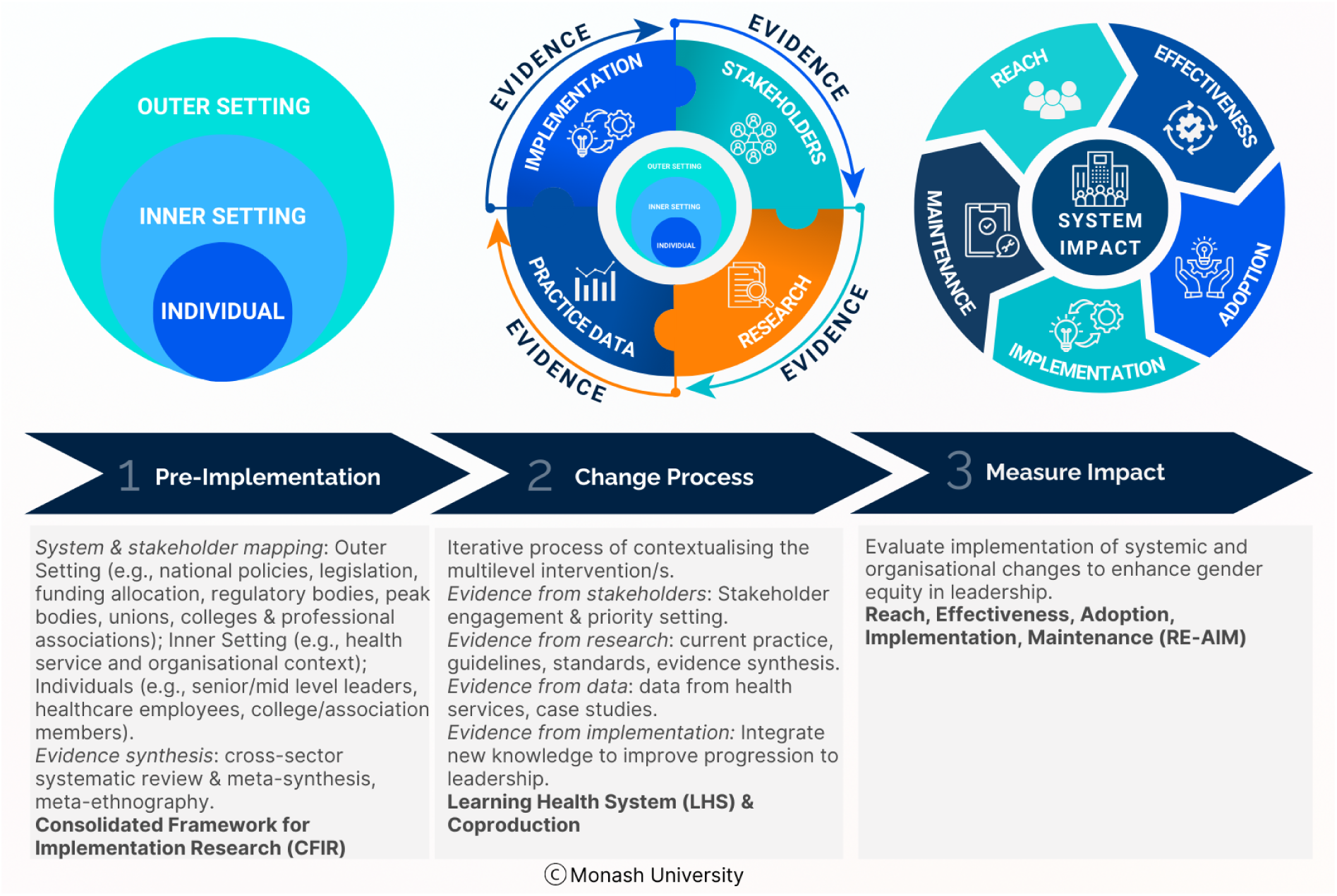
The Monash Systems Change Approach, incorporating the Consolidated Framework for Implementation Research (33), Learning Health System (38) and the Reach, Effectiveness, Adoption, Implementation and Maintenance (37) frameworks.

The CFIR will be used as a comprehensive ‘pre-implementation’ framework to systematically consider the roles, needs, and influences of diverse stakeholders. It supports the mapping of stakeholder groups and the contextualisation of gender equity strategies within the current system, while also informing the implementation and adoption of gender equity interventions across the system and organisations in healthcare (33). Spanning five core domains, CFIR comprises: 1) the outer context, e.g., government, policy, advocacy, and peak health bodies; 2) the inner organisational context, e.g., the norms, assumptions, culture, values and readiness of health services and academic centres; 3) the individuals in the system, e.g., women in the workforce such as clinicians, nurses, allied health staff, and management; 4) the evidence-based multi-level interventions to be implemented; and 5) the process of implementation.

While CFIR offers a robust framework for mapping systems and stakeholders, it functions primarily as a static, determinant framework. This limits its utility in complex systems where contextual factors can shift and evolve over time. CFIR is well-suited to identifying *what* multi-level factors are important (e.g., across the outer setting, inner setting, and individual levels), but it provides limited guidance on *how* to leverage these insights to coproduce context-sensitive strategies (34). To address this gap, we will apply the LHS framework, which supports iterative learning, stakeholder engagement, and adaptive implementation within complex and evolving systems (35,36). The LHS captures, generates and leverages evidence across four broad quadrants, including from stakeholder, research, practice/data and implementation to drive evidence into practice and towards systems and individual behaviour change, in this case for gender equity interventions in health leadership. The CFIR and LHS frameworks are complementary and will be applied to synergise research and implementation efforts.

The RE-AIM framework will be applied to evaluate the impact of systemic and organisational changes to enhance gender equality in leadership (37).

### Governance, partner organisations, settings and stakeholders

Led by the Monash Centre for Health Research and Implementation (MCHRI), Monash University, Australia, nine organisations formally partnered to establish AWHL, supported by two academic institutions from the United Kingdom. Partner funding was provided in cash and in-kind contributions, underpinning a successful competitive National Health and Medical Research Council (NHMRC) Partnership Grant. Following this, additional stakeholder partners and another academic institution joined the initiative with extended research themes, leading to a second successful competitive NHMRC Partnership Grant and total funding exceeding $6 million AUD.

Partnerships across leading health networks and professional medical and nursing colleges, government, and academia are now well established, supported by legal multi-institutional agreements, robust governance structures, and shared staff and resources.

Stakeholders and academics with expertise across health, business, and leadership established the project’s leadership and governance structure, including a steering committee to oversee research, implementation and evaluation. An advisory committee was formed to provide strategic guidance and expert advice. A project management team – including the authors – has also been established, bringing expertise in health, leadership, gender equity, and mixed-methods research to conduct the project and partner with stakeholders. The overall governance structure is illustrated in Figure 2.

**Figure 2.**
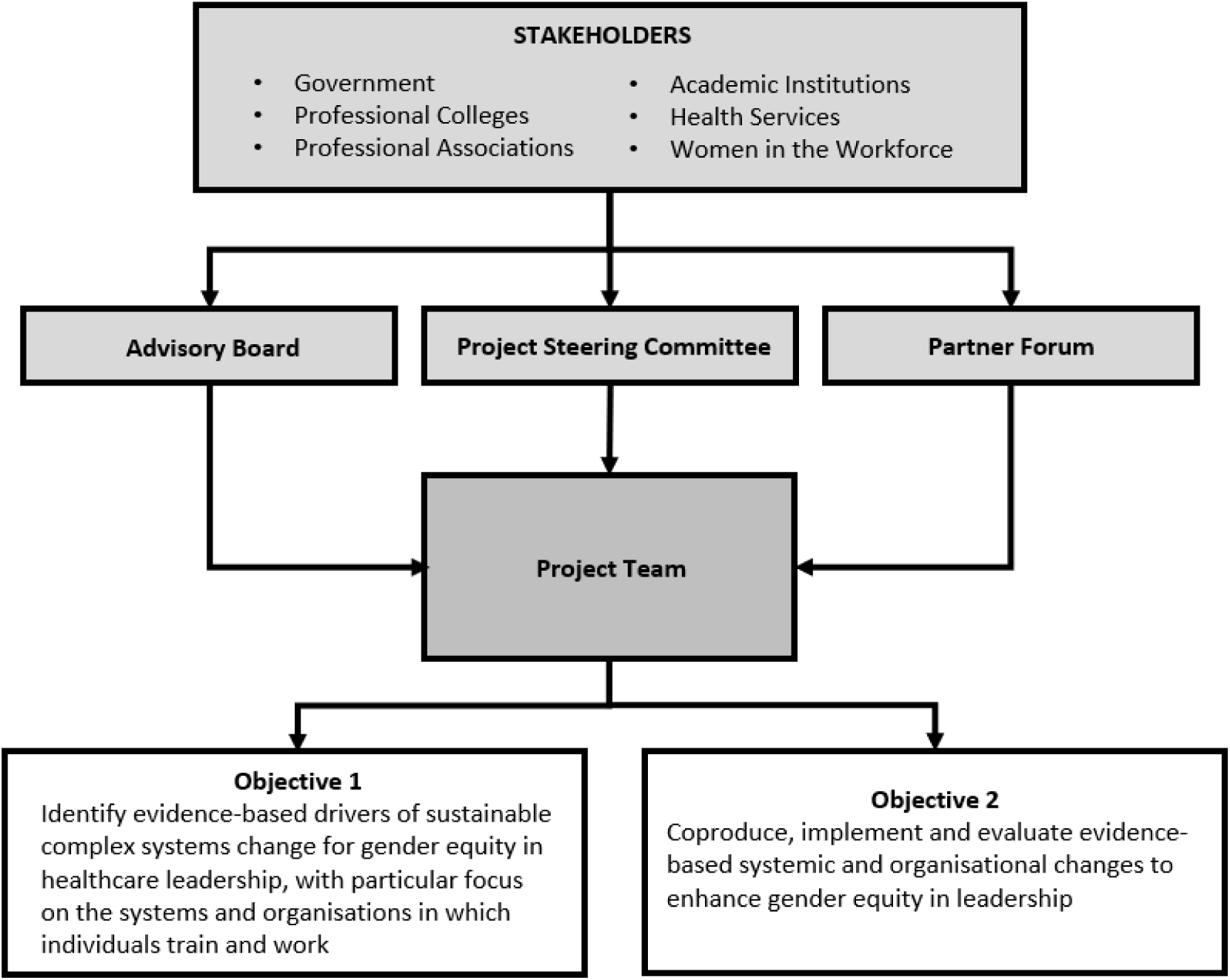
Overall OCM workstream governance structure.

AWHL directly engages with the stakeholder groups, brokered, supported and facilitated by academic researchers. Partner organisations include a broad range of stakeholders and settings; public and private health networks (across multiple hospitals and community health services), professional medical and nursing colleges, government agencies, and not-for-profit organisations, including one funded to support equity in the academic sector (Supplementary file 1). The workforce across these member organisations and healthcare networks constitutes a collective of women in the workforce recognised as key stakeholders. We will engage women in the workforce through surveys, qualitative interviews, and workshops, where they directly inform priorities. Engagement will also occur through women in representative roles (e.g., committees) at their organisations serving, as intermediaries between the project team and women in the workforce. A shared vision to advance women in healthcare leadership was developed through a codesign workshop in July 2021 with founding partners.

### Data collection tools and analysis

A range of data collection tools will be used (Figure 3). A systematic review and meta-synthesis (24), meta-ethnography (39), and modified grounded theory study (40) have been completed to identify: i) enablers and barriers to leadership capacity, capability and credibility; ii) effective interventions and strategies for organisational change to advance leadership for women; and iii) effective implementation strategies, theories and practical applications, including content and delivery methods.

**Figure 3.**
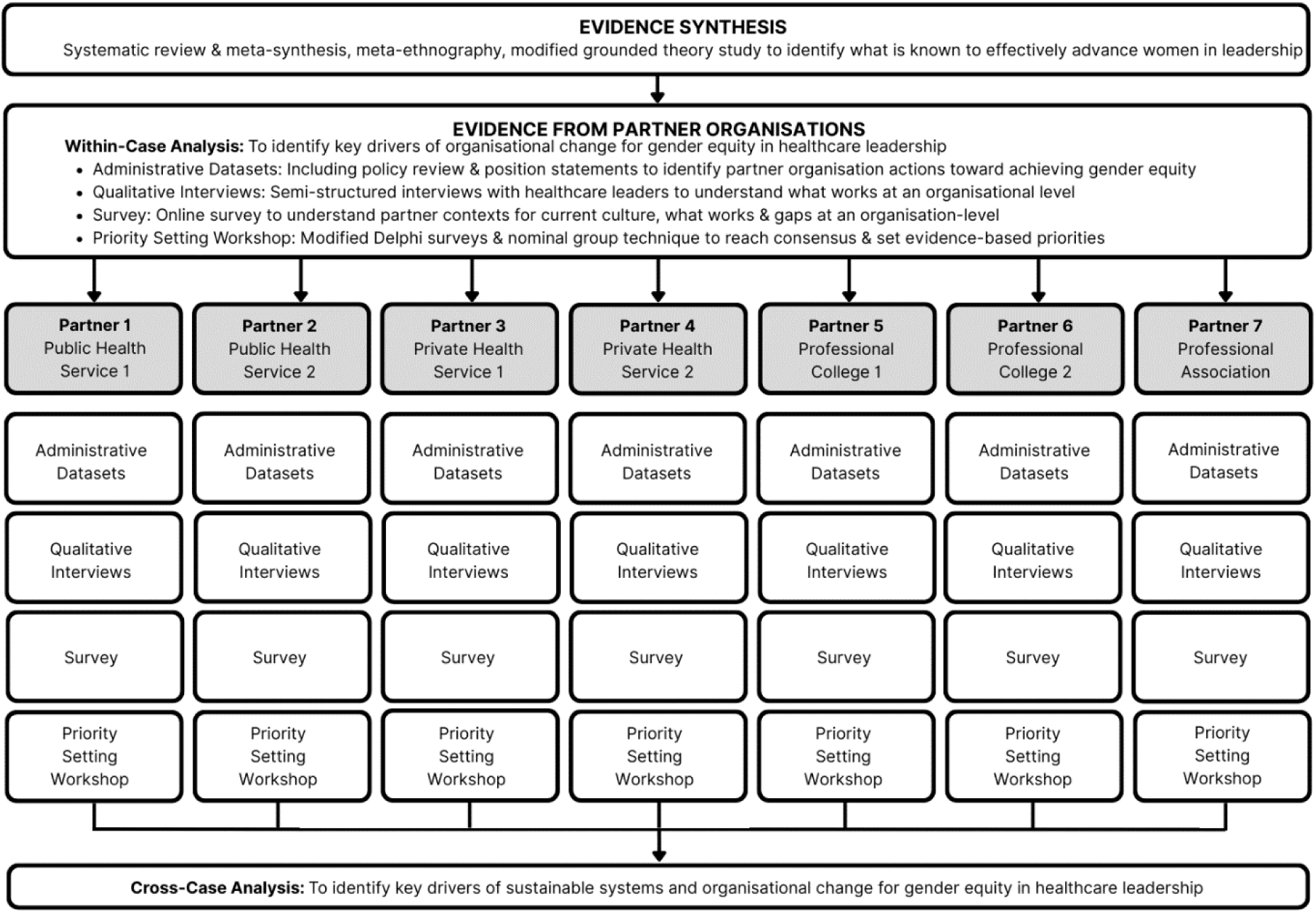
Process for data collection and analysis.

Further research with lead partners – each representing a distinct healthcare stakeholder context and constituting a single case (27) – will undergo in-depth exploration through purposeful engagement, to identify organisational strategies, strengths, gaps, opportunities and priorities for achieving gender equality in healthcare leadership. Aligned with our coproduction approach, lead partner organisations will be involved from the outset (26). Partners will work with the AWHL team to develop objectives, data collection tools, and participant recruitment strategies. This active input will continue throughout data collection, analysis, and dissemination to ensure shared findings and iterative improvements.

The three main data collection methods for each case are:

1. *Administrative datasets*. Partner organisation administrative datasets and policy documents will be reviewed to identify past and current initiatives aimed at promoting gender equity and equality in leadership. Information about gender representation across the workforce, training, and governance committees will also be examined.
2. *Semi-structured interviews.* Healthcare leaders will be interviewed to understand partner contexts and identify what works at an organisational level.
3. *Online survey.* Workplace employees and members of professional colleges and associations will be surveyed to understand partner contexts for current culture, identify what works and highlight gaps at an organisational level.

Interview guides and surveys will be co-designed, and questions informed by the Monash Systems Change Approach, evidence synthesis, and stakeholder feedback.

Semi-structured qualitative interviews will be undertaken with sample size guided by appraisal of information power – determined through appraisal of the study aim, sample specificity, use of theory, quality of interview dialogue and analysis strategy to develop new knowledge (41). Purposive sampling will be used across partners to identify women and men in leadership positions within targeted groups (e.g., across disciplines such as nursing, medicine, allied health; career stages; public and private sectors; culturally diverse groups; health service managers; and policy makers). All interviews will be audio-recorded, transcribed, and anonymised.

Surveys will be administered using Qualtrics software to health service employees and members of professional organisations. These instruments will generate both quantitative and qualitative data spanning career stages, disciplines, sectors (public and private), and individual versus organisational levels.

Analysis will first occur within each case (27). Template analysis will be utilised to thematically analyse qualitative data (42). Template analysis was chosen for its clear, systematic, and flexible approach. A coding template will be developed both inductively and deductively by the project team, guided by the study’s aim, evidence synthesis, Monash Systems Change Approach, while remaining open to new insights (42). The template will be finalised with input from partners and the project team, and the remaining dataset will then be coded to this template. Quantitative data will be descriptively analysed and summarised.

Findings from administrative datasets, interviews, and surveys will be triangulated within each case and fed back to individual partner organisations to support interpretation and collective sense-making, and identification of strengths, gaps and opportunities.

Once data collection and analysis are complete, each partner organisation will receive a report outlining their results, identifying what is working in their context and opportunity areas to advance women in leadership. A priority-setting workshop for each partner organisation will then be convened, led by the partner organisation and supported by the AWHL team. Modified Delphi surveys and nominal group techniques (43) will be employed to guide a consensus-driven identification of organisational priorities for tailored interventions. The priority setting process will comprise the following steps:

1. *Participant selection.* Partner organisations will identify and invite leaders and employees or members from across their organisations, ensuring diversity across career stage and gender.
2. *Part One: Pre-workshop ranking survey.* Two weeks prior to the workshop, all employees or members, including workshop participants, will be invited to complete a confidential and anonymous online survey (‘round one’). They will be presented with a list of contextualised evidence-based interventions – identified through the process outlined in Figure 3 – aimed at delivering gender equity outcomes that advance women into leadership at the organisation. Participants will be asked to rank the interventions they consider most important and acceptable.
3. *Part Two: Priority Setting Workshop.* Facilitated by the AWHL team, a three to four-hour workshop will include:

a. Overview of the evidence-base, context, and purpose.
b. Open discussion of the Top 10 ranked interventions from the pre-workshop survey.
c. ‘Round two’: Individual, confidential polling to rank the Top 10 interventions by importance.
d. Presentation of round two findings to participants.
e. Introduction of a feasibility framework and discussion of feasibility for the interventions ranked in point c).
f. ‘Round three’: Individual, confidential polling to rank interventions by feasibility.
g. Presentation of collectively agreed priorities to participants.
4. *Action Plan*. The agreed priorities will inform an Action Plan and an implementation and evaluation strategy for each partner organisation to deliver gender equity outcomes. The AWHL team will provide implementation support throughout.

Following the workshop, a report will be compiled and provided to the partner organisation for formal endorsement by its Executive and/or Board. Once context-specific data collection, analysis, and priority setting have been completed for each partner organisation, a cross-case analysis will be undertaken to identify shared learnings across all partners.

### Methodology

The methodology for the OCM workstream is grounded in implementation science and systems change theories, as outlined in Figure 4 and discussed in detail below.

**Figure 4.**
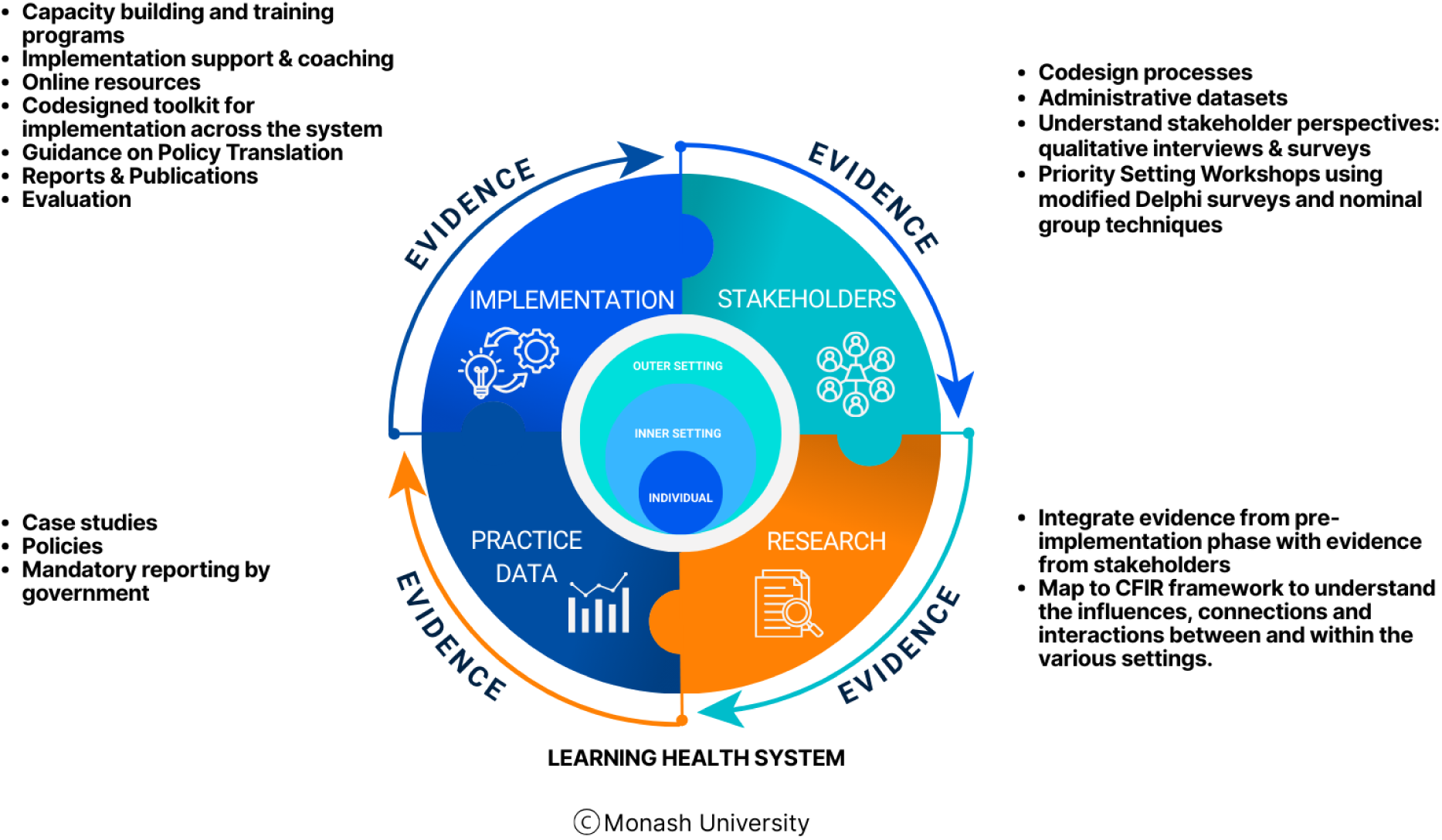
Methodology underpinning the project mapped against the Learning Health System. (36)

**Figure 5.**
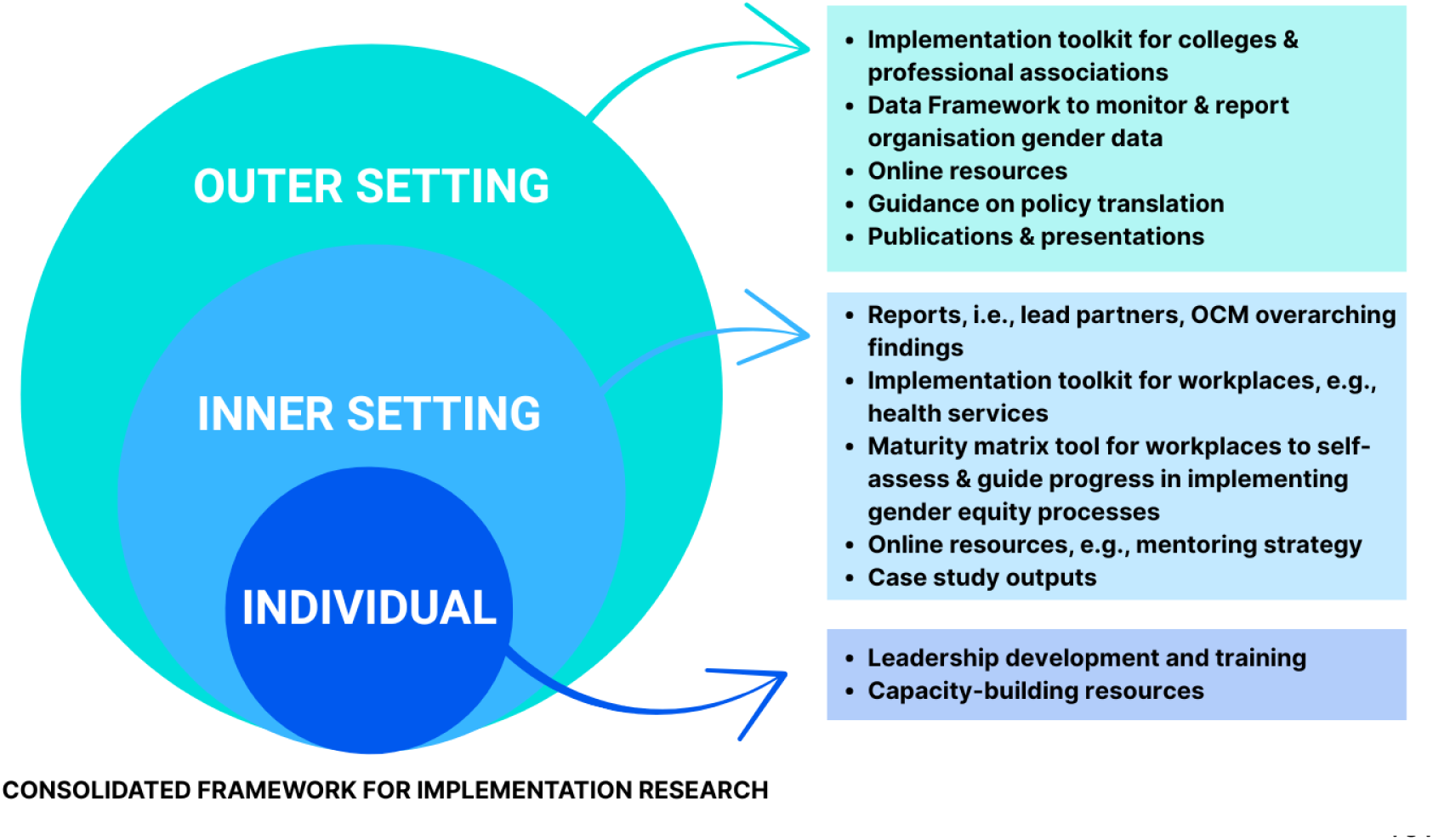
Implementation resource outputs mapped against the Consolidated Framework for Implementation Research. (33)

### Pre-implementation phase (Aim 1)

The aim of this phase is to identify evidence-based drivers of sustainable, complex systems change for gender equity in healthcare leadership, with particular focus on the systems and organisations where individuals train and work.

First, comprehensive system mapping activities will be undertaken to engage key stakeholders across both the outer and inner settings, and to maximise engagement and impact across systems levels (33). This process will identify stakeholders and clarify their roles – an essential step for fostering meaningful engagement and driving organisational change.

Second, evidence synthesis will assemble global and cross-sectoral evidence and map past and current gender equity initiatives to help identify the characteristics of multi-level interventions targeting organisational and systems change.

### Change phase (Aim 2)

The aim of this phase is to implement and evaluate evidence-based systemic and organisational changes to enhance gender equity in leadership. The change phase will be guided by the LHS framework, underpinned by coproduction, and will facilitate the process for iterative contextualisation, adaptation, implementation and evaluation of multilevel interventions. The LHS framework supports continuous learning cycles through integration of evidence from four interrelated quadrants: (i) stakeholder engagement and priority-setting, (ii) evidence synthesis and knowledge generation, (iii) current practice and data and (iv) implementation.

#### Quadrant 1: Stakeholder engagement and priority-setting

The first quadrant of the LHS framework, ‘Stakeholder derived evidence’, emphasises priority-setting and understanding stakeholder perspectives to ensure that developed interventions are relevant and responsive to their needs (36). Across each lead partner –representing distinct healthcare stakeholder contexts – administrative datasets will be analysed to identify past and current organisational initiatives aimed at promoting gender equity and equality in leadership. Subsequently, interviews with healthcare leaders (approximately 20 per partner) and surveys with employees or members will be conducted and analysed, as previously outlined. Data will be analysed individually for each partner and presented in tailored reports. To support strategic planning, priority setting workshops will be conducted within each lead partner organisation, facilitated by the AWHL project team. The aim of these workshops is to establish agreed, evidence-based priorities for delivering gender equity outcomes within each partner organisation. Modified Delphi surveys and nominal group techniques (43) will be used to identify consensus-based organisational priorities for advancing gender equity in leadership. Regular meetings with lead partners will be held to present and discuss emerging findings. Oversight will be provided by an established steering committee comprising delegates from each lead organisation.

#### Quadrant 2: Evidence synthesis and knowledge generation

The second quadrant of the LHS framework, ‘Research derived evidence’, focuses on generating knowledge and synthesising findings with existing research (36). To understand structures and processes required to drive, implement and evaluate systemic and organisational changes to improve gender equity in leadership, evidence synthesis from published literature and knowledge generated from the data collection methods described earlier (administrative datasets, interviews, surveys) will be mapped to the CFIR framework. This mapping will support understanding of the influences, connections, and interactions between and within the various settings.

#### Quadrant 3: Current practice and data

In the third quadrant of the LHS framework, ‘Data derived evidence’, emphasises understanding and integrating current practice and data to support iterative change processes (36). Research case studies will be informed by the findings outlined earlier. The implementation science and CFIR approaches address systems, organisational and individual perspectives, and hence enable exploration and evidence generation on systems change (e.g., across government, legislation and accreditation standards) and organisations (e.g., who create the workplace conditions and culture that can enable gender equity in leadership). Implementation strategies and resources will be developed to reflect findings and recommendations from semi-structured interviews and surveys that capture current practice in healthcare.

#### Quadrant 4: Implementation and healthcare leadership improvement

The fourth quadrant of the LHS, ‘Implementation evidence’, focuses on translating rigorous implementation research into practice to advance gender equity in healthcare leadership (36). Key outputs will include a codesigned implementation toolkit, developed using the UK Design Council Double Diamond codesign process. This process fosters codesign through four phases: (i) discover – working with stakeholders to understand the problem; (ii) define – focusing on specific problem areas; (iii) develop – identifying solutions with stakeholders; and (iv) deliver – testing and evaluating solutions (44).

Additional outputs will include capacity-building initiatives and policy translation guidance. Multi-layered strategies will be mapped to the CFIR framework to enable a targeted approach across each level of the system. Understanding the role of key stakeholders will provide insight into the support and changes required to promote adoption and drive meaningful change, as illustrated in Figure 4.

Building on this foundation, the next phase will focus on evaluating the effectiveness and sustainability of these strategies to ensure they translate into meaningful, system-wide change. Evaluation of the implementation toolkit and the multi-layered strategies, designed to support equitable and fair career pathways to leadership, will identify areas for refinement, thereby continuing the LHS cycle.

Implementation and evaluation will be codesigned with partner organisations. Evaluation will be guided by the RE-AIM framework, applied across reach (tool use and uptake), effectiveness (outcomes to be co-designed), adoption (integration into systems and organisational processes), implementation (fidelity of evidence-based processes and tools) and maintenance of outputs over time.

### Ensuring study quality

The AWHL initiative has undergone independent scientific peer review by the NHMRC under two successful Partnership Project Grants. Study quality and risk management will be monitored and ensured throughout the project by the steering committee and advisory board. The AWHL steering committee meets at least quarterly and provides guidance and advice to the project team to ensure all goals and objectives are met. The advisory board meets biannually and provides ongoing strategic guidance and expert advice to the steering committee and project team. This includes support in developing strong collaborative links, nationally and internationally, with relevant services, community organisations, government agencies, universities, and research centres.

The established national partnership, spanning professional medical and nursing colleges, professional associations, leading health services, and government, is guided by cross-sector academic expertise. Together with codesign and coproduction approaches, this structure reduces the risk of low organisational engagement and limited participant recruitment.

AWHL’s work is underpinned by robust methodology, including codesign and coproduction approaches (26), evidence synthesis (24,39,40), and CFIR and RE-AIM frameworks (33,37).

### Ethics and dissemination

Ethics approval was obtained from the Monash University Human Research Ethics Committee (Project ID: 25097). Findings will be disseminated through a variety of methods to maximise reach and engagement. This will include presentations of preliminary and final findings to individual stakeholder partners and their nominated committees and Boards, as well as presentations at national and international conferences. Manuscripts will be prepared for publication in open-access, peer-reviewed scientific journals to ensure the findings are rigorously presented and accessible to all.

## DISCUSSION

Achieving gender equity in healthcare leadership requires navigating a highly complex and dynamic system. Implementing change within such systems presents significant challenges, particularly when aiming for sustainable and embedded transformation. The OCM workstream provides the strategic mechanism for aligning change efforts across national policy frameworks, organisational structures, and the everyday practices that shape leadership opportunities (45,46).

Progress depends on alignment across multiple levels—from national policy frameworks to organisational cultures and structures that shape leadership pathways. To accelerate gender equity in healthcare leadership, the AWHL initiative adopts a multi-level, coproduced approach, ensuring that stakeholders across all levels of the system are actively engaged in shaping and delivering change (26,33). Stakeholders will partner with the AWHL team to design, implement, evaluate, and scale interventions at the system, organisational, and individual levels. These interventions aim to measurably improve career progression for women in healthcare leadership.

The methodology integrates robust implementation science frameworks, codesign principles, and comprehensive data collection strategies (26,33,36). At the systems level, both the outer setting—including national policies, legislation, funding mechanisms, regulatory bodies, unions, professional associations, and peak organisations—and the inner setting, such as health services and individual organisations, are considered. Interventions may include policy reform, awareness campaigns, support tools (e.g., measurement and evaluation frameworks), and educational opportunities, all designed to foster meaningful and lasting change for individuals within the system.

Without adequate support, strategic alignment, and leadership commitment, gender equity in healthcare leadership is unlikely to be achieved or sustained. Whole-of-system thinking is essential to drive meaningful progress. This requires consolidated efforts, prioritisation, resource allocation, and ongoing leadership commitment (24). Optimising implementation demands evidence on the “who,” “what,” and “how,” as articulated through the LHS quadrants. This protocol outlines a national, systems-level initiative that responds to policy and stakeholder priorities by addressing: “who” (stakeholders across outer, inner, and individual levels), “what” (implementation toolkits, resources, and support mechanisms), and “how” (policies, processes, and strategies tailored to each level).

To our knowledge, this is the first national partnership of its kind and scale—bringing together strategic partners and key stakeholders, including women in the healthcare workforce, professional colleges and associations, leading health services, government agencies, and cross-sector academic experts. This partnership aims to enable coordinated action to understand, develop, and implement sustainable, evidence-informed system and organisational change to advance women in healthcare leadership.

Our approach applies rigorous organisational change management methodologies, underpinned by a shared vision, coproduction, and a focus on measurable outcomes and impact. Across all phases, we aim to fast-track the translation of sustainable organisational change for broad scale-up and impact. By leveraging expertise in implementation science, scale-up strategies, behavioural science, and systems change, we present a novel framework for supporting system-level transformation—applicable not only to gender equity initiatives but to broader, complex equity and leadership challenges in healthcare.

## Data Availability

No datasets were generated or analysed for this protocol. The data arising from the study described in this protocol will not be shared, as it contains personal narratives that may reveal participant identities even after anonymisation.

## Abbreviations

OCM: Organisational Change Management
AWHL: Advancing Women in Healthcare Leadership
CFIR: Consolidated Framework for Implementation Research
LHS: Learning Health System
RE-AIM: Reach, Effectiveness, Adoption, Implementation and Maintenance framework
MCHRI: Monash Centre for Health Research and Implementation
NHMRC: National Health and Medical Research Council

## Acknowledgements

We would like to acknowledge and thank our lead partner organisations for their contribution. We also thank all members of the steering committee and advisory board for their support, enthusiasm and contribution to the project.

## Author contributions

HT, BG, JP, DR, EL, ES, GC, KR and MM conceptualised the original idea for the project and contributed to the development of the research protocol. BG and HT prepared the first draft of the manuscript, with ARC contributing to subsequent revisions. All authors contributed to critical revisions of the manuscript. All authors have read and approved the final version of the manuscript for submission.

## DECLARATIONS

Ethics approval was obtained from Monash University Human Research Ethics Committee (Project ID: 25097). Information about the project will be provided to, and consent obtained from, all participants completing interviews and online surveys.

## Consent for publications

Not applicable.

